# Assessment of the knowledge, preferences and concern regarding the prospective COVID- 19 vaccine among adults residing in New Delhi, India-A cross sectional study

**DOI:** 10.1101/2021.01.23.21250164

**Authors:** Farzana Islam, Rashmi Agarwalla, Meely Panda, Yasir Alvi, Vishal Singh, Arup Debroy, Arindam Ray, Amruta Vadnerkar, Shraddha Uttekar

**Author notes:** Corresponding author Dr Yasir Alvi, Department of Community Medicine, Hamdard Institute of Medical Sciences and Research, New Delhi.

## Abstract

**Background:** Understanding the perception and concerns of people about COVID-19 vaccine in developing and populous country like India will help in understanding demand for the vaccine and further tailoring out public health information and education activities before the launch of the vaccine. The study was carried out to assess the present state of knowledge people have about the probable vaccine for COVID-19, to know the preferences of respondents about this vaccine and to learn the expectations and apprehensions of people about features of this prospective COVID-19 vaccine residing in the capital city of India.

**Methods:** This cross-sectional study was conducted amongst the residents of Delhi, India from July-October 2020. Both offline and online interview method was used to collect date from 513 participants representing various occupational strata. Data was collected on socio demographic variable, vaccine acceptance and concerns regarding COVID-19 vaccine.

**Results:** Among the study population 79.5% said they will take the vaccine while 8.8% said they were not going to take the vaccine and remaining 11.7% had not yet decided about it. Most of them(78.8%),believed that vaccine would be available to public next year but at the same time half(50.1%) of them believe that it may not be in sufficient amount for everyone to get. More than 50% were willing to pay for the vaccine and 72% felt vaccine should first be given to health workers and high risk group.

**Conclusion:** The following study has helped to understand the percentage of people who are hesitant to take the vaccine and also the concerns regarding the vaccine. Also since half of the population is willing to pay for the vaccine, a strategical approach considering the various economical classes of people could be applied in a developing country like India.

## Introduction

COVID-19, as we know today is one of the biggest challenge mankind has faced in the century. The disease has jolted the entire world with more than 1 million deaths occurring worldwide. ^[1]^ India reported 88,74,290 cases and 1,30,519 deaths as on 17^th^ Nov 2020.^[2]^ There has been extensive global efforts to combat the pandemic. Efforts in resource constrained setting like India have also been multidimensional. ^[3]^ However, the disease has caused huge economic loss and has left the entire world socially crippled.

As mentioned by various epidemiologists the virus is here to stay. In such situations acquiring immunity against the virus becomes essential. Immunity can be acquired in two ways, either naturally or through vaccine. In the current pandemic the role of safe and effective COVID-19 will be very vital in the fight against COVID-19.Progress and claims about the vaccine trials have been mooted by several epidemiologist and medical experts, yet it is that opportunity which mankind has nailed all his hope upon. ^[4]^ The present pandemic situation warrants a potential accelerated timeline for development of vaccine with overlapping of phases and emergency use authorization. ^[4]^ Till October end two vaccines have been approved. Also, the Moderna, m RNA – 1273 and the AstraZeneca/Oxford – ChAdOxl nCoV-19 have already entered the phase III. India’s candidate vaccines COVAXIN and ZyCov-D which are in the pathway of progress to be able to deliver us immunity have moved over to phase II/III trials. Preliminary findings show a vaccine developed by Pfizer and BioNTech can prevent more than 90% of people from getting Covid-19 and Moderna vaccine has claimed 94.5% efficacy in its interim analysis.^[5]^ WHO estimates a total cost of 8 billion USD to develop a suite of three or more vaccines having different technologies and distribution to prevent COVID – 19 worldwide. ^[6]^ However, the efficacy, frequency, safety, preferences and precautions are some such queries that every individual whoever counts on the hope of vaccine advent, harbours in his mind.

Advent of any vaccine confronts plenty of issues. Based on prior studies about advent of other vaccines, we could enumerate that hurdles occur at the level of Vaccine – like production, manufacture, trials, marketing, safety and cold chain maintenance. Ethics and efficacy or beneficiary – like acceptance, fear, compliance, willingness to pay, site predilection or an apprehension for any adverse effects. Vaccine acceptance is a social tool which plays an important role for advent, implementation and continuation of any vaccination programme. ^[7,8]^ Accounting for the perception of people about vaccine advent will give empirical details about their aspirations and apprehensions. Considering factors like willingness to pay will help in considering the financial constraints and economies into consideration before a large scale bulk production. Decisions like whom to administer first, where and how to administer, cost constraints, willingness to come out to health facilities for getting the shots or volunteering to be part of such trials will further ease the policy and programmatic commitments. Thus this study is being conducted with the objectives to assess the present state of knowledge people have about the probable vaccine for COVID-19, to know the preferences of respondents about this vaccine and to learn the expectations and apprehensions of people about features of this prospective COVID-19 vaccine residing in the capital city of India.

## Materials and Methods

### Study design, setting and participants

The cross-sectional quantitative study was carried out among the residents of Delhi, North India from July-October 2020.

Various sections of society were taken.

➢ Occupation such as: All staff in health work force including doctors, nurses, paramedics and health workers
➢ Staff in corporate sectors including engineers, technicians, managers and leaders etc
➢ Business class including Shopkeepers, owners, delivery boys, Managers etc…
➢ Education sector – who constitute a major chunk and play major role in taking care of young ones like teachers, principal and tutors etc.

### Sample size

In such a dynamic scenario where figures escalate and slump on a day to day basis, considering the rule of assumptions a prevalence of 50 % was considered and Schwartz formula was applied. At 90% power and 95% confidence interval, the sample size came out to be 400. Considering a non-response rate of 10 %, sample size of 440 was obtained and further rounded of to 500.A total of 513 participants responded.

### Procedure

Respondents above 18 years of age, who consented, were included in the study, which was either conducted physically as an interview or using online technique, depending on the respondent’s convenience.

For the physical interview method, individuals were selected by simple random sampling from among these strata’s so as to reach our estimated sample size. After a brief training, the interviewer collected data from these subjects by a semi structured questionnaire which was pre-tested and expert validated.

### Online method

For those individuals who consented but had time constraint, we wilfully administered the questionnaire to them in Microsoft forms and awaited response from them via the link provided. The terms not understood by them were clarified and explained to them over phone. All terms of ethical consideration, beneficence, autonomy and confidentiality was followed.

The questionnaire was semi structured and contained the socio demographic details, respondent’s perception and knowledge about the prospective COVID-19 vaccine, questions regarding their expectations, fears, barriers and apprehensions about such a vaccine. The last part included questions regarding their promptness to be part of any such vaccine trials for the benefit of mankind.

The questions were validated by experts and repeat consultations and modifications done as required after an initial pilot study. A link was created to be able to reach professionals who consented to be part of the study but owing to time constraint preferred to respond to it as per their own convenience. **https://forms.office.com/Pages/ResponsePage.aspx?id=DQSIkWdsW0yxEjajBLZtrQAAAAAAAAAAAAN__swYqWBUQkwyV09QMDEyOEwxME8zUVpUQTgxMUI1UC4u** The preference of individuals for Vaccine in the terms of its availability, efficacy, intake and cost; mounted on a likelihood scale with 1 depicting least likely and 6 depicting most likely was taken up as the dependant variables. Socio-demographic details like age, caste, job profile, knowledge about the vaccine’s safety, efficacy, time of advent, adequacy etc., the willingness to be part of trial, preferences for route of administration, readiness to spend and take booster doses were also analysed.

### Data analysis

The completed questionnaire was checked for completeness and consistency. Collected data was entered in the MS Excel spreadsheet, coded appropriately and later cleaned for any possible errors in a SPSS (Statistical Package for Social Studies) for windows version 21.0. Categorical data was presented as percentage (%). Pearson’s chi square test was used to evaluate differences between groups for categorized variables. In case, the expected cell count was found to be less than 5 in >20% cells, we used the fisher’s exact test. Normally distributed data was presented as means and standard deviation with 95% confidence intervals (CI). All tests were performed at a 5% level significance, and thus the value less than 0.05 (p value < 0.05) was taken as significant association.

### Ethics statement

Permission to conduct the study was taken from the Research Proposal Advisory Committee and approval was taken from the Institutional Ethics Committee. Informed consent in writing was taken from respondents who participated through physical interview and online consent was taken by participants who filled the form online. There were no conflict of interest and all measures of autonomy and confidentiality were strictly maintained.

## Results

A total of 513 participants responded. Amongst all the participants, almost equal participation were there from male (50.9%) and females (48.3%). Most of the participants were middle aged (41.7%), Hindu by religion (65.9%) and currently married (57.7%), Most of the study participants (88.5%) were either in college or had completed college. Among occupation of participants, most of them were health care workers like doctor/ nurse/ paramedic/ ANM (42.7%), 23.2% were students, and rest were from other service sectors.

Based on the history of COVID-19 infection among the study participants, 2.5% had COVID-19 infection in the past, and 1.4% said there was mortality due to COVID-19 in their family. Around three fourth had a moderate to strong feeling that they may catch the infection anytime, while almost two third of those who had COVID-19 infection in past, think that re infection can happen. Most of them were afraid of dying after getting COVID-19. **(Table 1)**

**Table 1:** COVID-19 related experiences and perceptions among study population

| Variables | Frequency | Percentage |
| --- | --- | --- |
| **Did you have COVID-19 in the past** |  |  |
| Yes | 13 | 2.5 |
| No | 500 | 97.5 |
| **Did anyone in family COVID-19 in the past** |  |  |
| One person | 26 | 5.1 |
| More than one person | 23 | 4.5 |
| None | 464 | 90.4 |
| **Did anyone in family died due to COVID-19** |  |  |
| Yes | 7 | 1.4 |
| No | 42 | 8.2 |
| No COVID in family | 464 | 90.4 |
| **How likely do you feel that you can get COVID-19 anytime<sup>@</sup>** |  |  |
| Not at all | 26 | 5.3 |
| Some time | 109 | 22.0 |
| Moderately feeling | 295 | 59.6 |
| Strong feeling | 65 | 13.1 |
| **How likely do you feel you can get COVID-19 reinfection<sup>#</sup>** |  |  |
| Not at all | 2 | 15.4 |
| Some time | 3 | 23.1 |
| Moderately feeling | 4 | 30.8 |
| Strong feeling | 4 | 30.8 |
| **How likely do you feel that you may die from COVID-19<sup>^</sup>** |  |  |
| Not at all | 90 | 17.8 |
| Some time | 271 | 53.5 |
| Moderately feeling | 132 | 26.0 |
| Strong feeling | 14 | 2.8 |

<sup>@</sup> total 495
<sup>#</sup> Total 13 (If previously infected)
<sup>^</sup> Total 507

**Figure 1** shows COVID-19 vaccine acceptance. It was observed that 79.5% would take the vaccine whenever available, while 8.8% said they would turn it down and remaining 11.7% had not yet decided about it. **Figure 2** shows knowledge and attitude of study population about the COVID-19 vaccine. Most of them believed that vaccine would be available to public next year (78.8%), but at the same time half of them believe that it may not be in sufficient amount for everyone to get (50.1%) and 52.4% did feel it will not prevent the COVID-19 disease for lifelong. Majority knew that the COVID-10 vaccine would not prevent all types of pneumonia (67.4%).

As per **Table 2**, the study shows that participants were willing to take COVID-19 vaccine by any route in any number of doses, but preferably with a good effectiveness and rare side adverse reaction. People were willing to get the vaccine even if they need to pay for it (84.4%), and had to get vaccinated every year (75.6%).

**Table 2:** Vaccine related preferences among study population

| Variables | Frequency<br>n=513 | Percentage |
| --- | --- | --- |
| **Preferred route** |  |  |
| Any route | 207 | 40.4 |
| Any form of injection | 152 | 29.6 |
| Oral drops/ jet | 104 | 20.3 |
| Inhalation by nose | 42 | 8.2 |
| Skin patch | 8 | 1.6 |
| **Preferred dose** |  |  |
| Any number of doses | 167 | 32.6 |
| Four doses | 30 | 5.8 |
| More than four doses, but less than seven | 21 | 4.1 |
| Three doses | 165 | 32.2 |
| Two doses | 130 | 25.3 |
| **Minimum acceptable effectiveness** |  |  |
| More than 90% | 122 | 23.8 |
| 80-90% | 199 | 38.8 |
| 60-79% | 148 | 28.8 |
| 40-59% | 35 | 6.8 |
| Even below 40% | 9 | 1.8 |
| **Least tolerable side effect for you** |  |  |
| Fainting | 206 | 40.2 |
| Fatigue | 32 | 6.2 |
| Fever | 49 | 9.6 |
| Injection site Pain | 131 | 25.5 |
| Rash | 54 | 10.5 |
| Sleepiness | 41 | 8.0 |
| **Acceptable adverse reaction** |  |  |
| No risk is acceptable to me | 89 | 17.3 |
| Worse disease 1 in 100 vaccinated | 17 | 3.3 |
| Worse disease 1 in 1000 vaccinated | 19 | 3.7 |
| Worse disease 1 in 1000 vaccinated | 50 | 9.7 |
| Worse disease 1 in 1 lakh vaccinated | 141 | 27.5 |
| Worse disease 1 in 10 lakhs vaccinated | 197 | 38.1 |
| **Preferred price for COVID vaccine** |  |  |
| Will accept if it's free | 80 | 15.6 |
| 100-500 INR | 197 | 38.4 |
| 500-2000 INR | 188 | 36.6 |
| Will take even if >2000 INR | 48 | 9.4 |
| **Preferred place where you like to get it** |  |  |
| Anywhere | 90 | 17.5 |
| From any health facility | 276 | 53.8 |
| From our house physician | 45 | 8.8 |
| Through campaign | 41 | 8.0 |
| At our home, by a trained person | 42 | 8.2 |
| Self-administration | 19 | 3.7 |
| **If required, will you take the vaccine every year** |  |  |
| No | 125 | 24.4 |
| Yes | 388 | 75.6 |
| **Have you taken yearly flu vaccine** |  |  |
| Never | 316 | 61.6 |
| Once | 81 | 15.8 |
| Occasionally | 91 | 17.7 |
| Regularly, each year | 25 | 4.9 |
| **Is your child immunized** |  |  |
| Yes, fully vaccinated as per age | 297 | 57.9 |
| Missed some doses | 12 | 2.3 |
| No, don't believe in vaccines | 3 | .6 |
| I have no kid | 201 | 39.2 |
| **Would you volunteer for Human Challenge study** |  |  |
| No | 316 | 61.6 |
| Yes | 197 | 38.4 |
| **Who should get the vaccine first** |  |  |
| Health care workers | 261 | 50.9 |
| Elderly | 48 | 9.4 |
| People with diseases like diabetes | 59 | 11.5 |
| Children less than five | 37 | 7.2 |
| Essential service | 27 | 5.3 |
| Everyone should get at same time | 81 | 15.8 |

**Table 3** shows associations of COVID-19 vaccine acceptance response with demographic factors. It was seen that lower age, females, Hindu by religion, Secondary or higher educated, currently not married and students would accept the vaccine most in their categories if offered to them. It was observed the participants who were of lower age, Hindu by religion, currently not married and students were significantly associated with being more acceptable to COVID-19 vaccine uptake.

**Table 3:** Associations of Vaccine acceptance response with socio-demographic factors

| Variables |  | Vaccine acceptance response |  |  | P value |
| --- | --- | --- | --- | --- | --- |
|  |  | Will accept | Not yet decided | Refuse |  |
| **Age** | <35 | 227 (84.7%) | 26 (9.7%) | 15 (5.6%) | F=14.93,<br>p= 0.004 |
|  | 35-54 | 160 (74.8%) | 26 (12.1%) | 28 (13.1%) |  |
|  | ≥55 | 21 (67.7%) | 8 (25.8%) | 2 (6.5%) |  |
| **Gender** | Male | 200 (76.6%) | 31 (11.9%) | 30 (11.5%) | F= 6.142,<br>p= 0.151 |
|  | Prefer not to say | 3 (75.0%) | 1 (25.0%) | 0 (0%) |  |
|  | Female | 205 (82.7%) | 28 (11.3%) | 15 (6.0%) |  |
| **Religion** | Hindu | 282 (83.4%) | 33 (9.8%) | 23 (6.8%) | F= 15.565,<br>p= 0.003 |
|  | Muslim | 102 (75.0%) | 16 (11.8%) | 18 (13.2%) |  |
|  | Others | 24 (61.5%) | 11 (28.2%) | 4 (10.3%) |  |
| **Education** | Illiterate | 4 (66.7%) | 1 (16.7%) | 1 (16.7%) | F= 6.214,<br>p= 0.296 |
|  | Primary | 5 (62.5%) | 3 (37.5%) | 0 (0.0%) |  |
|  | Secondary | 38 (84.4%) | 4 (8.9%) | 3 (6.7%) |  |
|  | College | 361 (79.5%) | 52 (11.5%) | 41 (9.0%) |  |
| **Marital status** | Married | 219 (74.0%) | 44 (14.9%) | 33 (11.1%) | $\chi^2= 13.220$,<br>p= 0.001 |
|  | Current not married | 189 (87.1%) | 16 (7.4%) | 12 (5.5%) |  |
| **Occupation** | Healthcare worker | 164 (74.9%) | 26 (11.9%) | 29 (13.2%) | $\chi^2= 20.925$,<br>p= 0.001 |
|  | Others | 107 (78.1%) | 18 (13.1%) | 12 (8.8%) |  |
|  | Student | 109 (91.6%) | 8 (6.7%) | 2 (1.7%) |  |
|  | Unemployed | 28 (73.7%) | 8 (21.1%) | 2 (1.7%) |  |

## Discussion

This study is first of its kind done in India with the objective to assess the knowledge, preferences, expectations and apprehensions about the probable vaccine for COVID-19 among residents of Delhi. It was observed that overall participants were quite aware about the COVID-19 vaccine and likely to get a vaccine if it were available.

In this unprecedented COVID-19 situation, some stress and fear among participants about catching the infection, being reinfected and concern of death due to the disease was also observed. The country wide lockdown, infodemics, etc. may have precipitated this stress and fear. About one-tenth of respondent had COVID-19 in their family, with mortality rate of 16.7%, which are higher than the national estimates. ^[9]^

Among the respondents, 79.5% were willing to accept the COVID-19 vaccine if it is available, 11.7% were not sure at the time of survey while 8.8% refused to take it. This acceptability rates in our study are higher in comparison to previous researches from other developing as well as developed countries. A study from Nigeria reported acceptability at 29%, in Saudi Arabia it was 65%, in Indonesia it varied between 67 – 93%, while in China it was 72.5% among the general population. ^[10, 11, 12, 13,]^ Among developed countries, a study in US reported 67% would accept a COVID-19 vaccine, while researchers in France documented about 25% of respondents would not use the vaccine when it is available.^[14,15,16]^ The scepticism were also reported in Larson’s Vaccine Confidence Project which documented one fifth of Swiss and 18% of French would refuse a COVID-19 vaccine.^[16]^ In a US poll, only half of the people said they will take a COVID-19 vaccine and 20% turned it down, while in a UK poll the refusal rate was about one in six.^[17,18]^ A multicentric study conducted in seven European countries found acceptance rate at 73.9%, while 7.2% desired not to get vaccinated.^[19]^ Another recent multicentric Ipsos survey, conducted on behalf of the World Economic Forum, observer overall three-quarters of adults agreed to get vaccinated when available. ^[20]^ In the same survey, India with 87% acceptance rate were among countries where COVID-19 vaccination intent is highest. ^[20]^ The higher acceptability in the study reflects good public confidence in COVID-19 vaccine. It may be due to many reasons which are seen in Indian subcontinent. First, the overall immunization coverage and vaccine acceptability in Indian is higher due efforts of government of India in collaboration with UNICEF and GAVI in providing free as well as universal vaccination. Secondly, the misinformation being spread by The Anti-Vaxx movement prevalent in developed countries are not so common in India. ^[21]^ While good acceptance rate in our study is depicts the positive attitudes towards a vaccine in India, we should also pay attention to portion who have not yet decided. These are the potential candidates for intervention for increasing vaccination acceptance among community.

In regards to factors effecting vaccine acceptance, it was observed the future COVID-19 vaccine acceptance was relatively higher among younger age group (84.7% among 34 or lower), females (82.7%), currently not married (87.1%), students (91.6) and participants with secondary education level (84.4%). Except gender and education level, all these associations were significant in statistical analysis. These finding were contrast to study from US and Saudi Arabia where they observed acceptance higher among older adults. ^[19, 20]^ Apart from different demography, social structure and political engagement, the vaccine acceptance was also different from our study, leading to different findings.

In the study, it was observed that people have mixed expectation from the research team working on COVID-19 vaccine candidates. As high as two-fifth of the respondents were expecting the vaccine to be released in India by end of this year for public use, while about 80% were expecting to come by next year. These rates were similar to the one observed in multicentric study where 59% disagreed that COVID-19vaccine would be available to them before the end of 2020. ^[20].^ While the Indian Council of Medical Research (ICMR) claims of early release have fuel this eagerness, it also creates doubts and pessimistic views of people about its efficacy and safety.^[22]^ This also reflects the desperation of the people to get vaccinated for COVID-19. Although the respondents are eagerly waiting for vaccine, they knew that it would be far from the perfect one. Most of them desired not to volunteer to participate in “human challenge” study which could accelerate its development. More than half were in the view that vaccine would provide lifelong immunity and it would not be sufficient for everyone when it will be available. About one-third said it would not protect all alike, while some of the respondents were searching alternatives to vaccine in homeopathy (12.3%) and Ayurveda (16.4%). Perception that vaccine would not be sufficient for everyone and may not protect all with lifelong immunity, could affect the vaccine acceptance rate in community. Although we observed handful proportion of sample searching alternatives to vaccine, a recent report by Wellcome Trust observed that almost all Indian (98%) agree that vaccines are important for children. ^[23]^

In regards to vaccine preferences, most of the participants were open to all the forms of routes, while about one third preferred injections over oral/inhalation. Almost one third were willing to have any number of doses, while other one-third were only willing to take a maximum number of three doses. The least accepted side effect for the participant was fainting (40%) and injection site pain (25%). This finding is important concern for vaccine candidate profile because it has been documented that most common reason for not getting covid-19 vaccine was concerned about potential side effects. ^[19,20]^ Almost two-fifth would immunize themselves only if the vaccine is at least 80-90% efficacious while rest one-fourth opt for at least 60% efficacy.

Similarly a study from Indonesia, reported 93.3% of respondents agreed for vaccination with 95% effective vaccine, but 67.0% accepted for a vaccine with 50% effectiveness. ^[24]^ Most of them were willing to take the vaccine every year, and would pay for it too. Almost same proportion of respondent were willing to pay100-500Rs, and between 500-2000Rs from their pocket for the vaccine, while about 16% would be vaccinated only if it was provided for free. These are similar to the findings from other developing country. ^[24]^ More than half of the respondents said that they like to get the COVID-19 vaccine from any health facility, which shows the community confidence in public health infrastructure, while confidence in vaccine was displayed by almost all the respondents who had children and had fully vaccinated them. Utilizing existing vaccine-distribution infrastructure is also preferred approach in planning COVID-19 vaccine distribution. Indians have good trust in doctors and health system as reported by recent report by Welcome Trust. ^[23]^ This is predominantly due to large numbers of community health workers which again build the confidence of community in the health system. This would be helpful in planning COVID-19 vaccination. Study from Saudi Arabia observed acceptance for COVID-19 vaccine was three times if they have trusted the health system. ^[13]^ The participants were aware that health care workers are the most at-risk population for getting COVID-19, more than half advocated that health care workers should get the COVID19 vaccines first, while there were few who said that the vaccine should be given to all at the same time. As a low-middle-income country with limited health budget, India cannot run the rat race of vaccine nationalism where developed countries are hedging their bets to secure potential vaccines supplies to their citizen, our focus should be on establishing a framework for the equitable allocation of Covid-19 vaccines including prioritizing high risk groups. ^[25]^

### Limitations

Only a sub sample of the population could be included in the study. Large scale studies from whole of India are needed to understand the knowledge, expectation and apprehension before the launch of the vaccine. Another limitation of the study was that it did not have equal representative from various economic and occupational strata of society which could bias the result.

## Conclusion

The study shows that COVID-19 vaccine hesitancy which is currently the concern in other countries is not an issue in India. Also with the launch of vaccine, vaccine misinfodemics will have to be tackled. As the country waits for the introduction of a vaccine against COVID-19, this pre-introduction period provides a great opportunity to develop a communication package to improve vaccine confidence in the community. This will help to improve the uptake of COVID-19 vaccine. This will also lead to overall immunization programme strengthening, with scientific evidences, clear and consistent communication and improved health literacy of both the community as well as the service providers

## Data Availability

The data that support the findings of this study are available from the corresponding author, upon reasonable request.

